# Laboratory-Developed Test Orders in an Academic Health System

**DOI:** 10.1101/2022.12.12.22283358

**Authors:** Jenna Rychert, Robert L. Schmidt, Jonathan R Genzen

**Affiliations:** University of Utah Health, Department of Pathology; ARUP Laboratories, Salt Lake City, UT; LabCorp, Burlington, NC

**Author notes:** <u>Corresponding Author</u> Jonathan R. Genzen, MD, PhD, ARUP Laboratories, 500 W Chipeta Way, mail code 100-G02, Salt Lake City, UT 84108.

## Abstract

**Importance:** The Verifying Accurate Leading-edge IVCT Development Act, if enacted, would create a unified regulatory oversight system for all in vitro clinical tests, including laboratory-developed tests.

**Objective:** To determine the frequency of use of laboratory-developed tests in an academic medical center system.

**Design:** Quality improvement study analyzing 2021 test order data.

**Setting:** Academic medical center (hospital, outpatient clinics, and cancer center) and non-profit national reference laboratory.

**Main Outcome(s) and Measure(s):** Main outcome, not applicable; non-interventional study of retrospective data. Measures include assay type, assay methodology, compliance status (i.e., Food and Drug Administration cleared, approved, and/or authorized assay, laboratory-developed test, and standard method), test order volume, inpatient versus outpatient setting, and provider medical specialty.

**Results:** Of the 3,016,928 tests ordered in 2021, 2,831,489 (93.9%) were Food and Drug Administration cleared, approved, and/or authorized assays, 116,583 (3.9%) were laboratory-developed tests, and 68,856 (2.3%) were standard methods. Laboratory-developed tests were more commonly ordered in the outpatient versus inpatient setting and represented a higher proportion of the test volume at the cancer center compared to University Hospital (5.6% vs 3.6% respectively). The top 167 laboratory-developed test assays accounted for 90% of the laboratory-developed test volume (104,996 orders). Among the 20 most frequently ordered laboratory-developed tests were mass spectrometry assays and tests used in the care of immunocompromised patients. Internal/family medicine placed the greatest number of orders (1,044,642) and ordered one of the lowest proportions of laboratory-developed tests (3.2%). Non-infectious disease molecular testing made up 8.8% of laboratory-developed tests ordered.

**Conclusions:** Laboratory-developed tests made up a small percentage of the total laboratory tests ordered within the academic health system studied. Regulatory reform proposals should consider the need for both safety and availability of laboratory-developed tests in clinical laboratory settings.

**KEY POINTS:** *Question:* How frequently are laboratory developed tests (LDTs) used in an academic medical center (AMC) setting?

*Findings:* In this quality improvement study looking at test orders in 2021, 93.9% of test orders were for FDA cleared, approved, or authorized assays, 3.9% were for LDTs, and 2.3% were for standard methods. The top 167 LDT assays accounted for 90% of the LTD volume.

*Meaning:* In vitro diagnostic reform efforts will impact many LDTs assays with relatively low order volumes in AMC settings.

## INTRODUCTION

The U.S. Food and Drug Administration (FDA) is authorized under the Medical Device Amendments of 1976 (MDA) to regulate medical devices, including in vitro diagnostics (IVDs), which are introduced into interstate commerce for commercial distribution.^1^ The MDA and subsequent federal regulations established the framework under which manufacturers of IVDs are required to obtain clearance or approval prior to distributing test kits or instruments used to diagnose human disease. FDA officials have described this as the “commercially distributed pathway” for IVD regulatory oversight.^2^

However, some clinical laboratory assays are developed within a single laboratory and are not distributed as kits for other laboratories to use. The FDA defines these laboratory-developed tests (LDTs) as “an IVD that is intended for clinical use and designed, manufactured, and used within a single clinical laboratory.”^3^ The FDA has asserted that it has the authority to regulate LDTs but that it has followed a policy of enforcement discretion.^3^ Although the FDA announced its intention to exercise oversight over LDTs in 2010 and released a draft regulatory framework in 2014, this framework was ultimately not implemented.^3-7^ The regulatory landscape surrounding LDTs remains controversial.^8-10^ Additionally, federal regulations from the Centers for Medicare and Medicaid Services (CMS) enacted under the Clinical Laboratory Improvement Amendments of 1988 (CLIA) have specified the performance standards for LDTs that clinical laboratories must follow prior to their use in high-complexity clinical laboratory settings.^11,12^

The Verifying Accurate Leading-edge IVCT Development Act (VALID Act) is a bill that was introduced into the U.S. Congress. If enacted, it would provide a unified regulatory oversight system for all in vitro clinical tests (IVCTs), a new definition that includes both IVDs and LDTs.^13,14^ A concern raised by the supporters of diagnostic reform is that the number of LDTs currently on the market and being used in patient care is not known.^15,16^ Clinical laboratories are specifically exempt from device registration with the FDA under current federal regulations.^17^ However, they do maintain and submit information about test menus to regulatory and accreditation agencies. For example, when applying for a CLIA certificate of compliance, a clinical laboratory must provide CMS with a full list of all assays performed, including their manufacturers.^18^ Further, laboratories accredited by the College of American Pathologists (CAP) are required to maintain a laboratory activity menu and a separate list of LDTs to assist in the process of biannual inspections, but this list is not submitted to the CMS.^19^ Finally, the New York State Department of Health (NYSDOH) maintains a database of all LDTs approved by the Wadsworth Center.^20^

As an academic health system with a university-owned national reference laboratory, our institution maintains information on all clinical laboratory testing performed within our facilities, including assay type, test volumes, and regulatory compliance status. As such, the present study was conducted to determine how frequently LDTs were ordered in inpatient and outpatient settings.

## METHODS

### Healthcare Setting

ARUP Laboratories is a not-for-profit enterprise of the University of Utah Department of Pathology. Along with serving as a national clinical reference laboratory, ARUP operates inpatient and outpatient clinical laboratories for the University of Utah Health, an academic medical center that includes the University Hospital, the Huntsman Cancer Institute, and more than a dozen community health centers and clinics.

### Sample

Throughout the manuscript, the term “assay” is used to refer to a distinct type of laboratory diagnostic method available in our test directory, whereas “test” refers to the performance of the specific assay method on a unique patient specimen under a clinician’s order. Under an Institutional Review Board exemption protocol (University of Utah, #00082990), a de-identified dataset was obtained for tests ordered by providers at the University of Utah Health from January 1 to December 31, 2021. The dataset included assay name, volume of test orders by year, patient admission status (inpatient and outpatient), location (University Hospital and Clinics [UH] and Huntsman Cancer Institute), and ordering department. Ordering departments were categorized according to the medical specialty. A subset of information on test orders collected at outpatient phlebotomy sites that could not be attributed to specific ordering departments in this dataset was categorized as unclassified. The dataset was then cross-referenced to the compliance status of the assay as an FDA assay (i.e., FDA cleared, approved, or exempt), emergency use authorization (EUA) assay, LDT (subclassified as in-house developed, analyte-specific reagent, or modified FDA), or standard assay.^21^ The classification of standard methods adhered to the NYSDOH definition of “a standardized protocol that is universally applied in laboratories that employ the method for the analyte” (e.g., immunohistochemical stains and in situ hybridization probes with pathologist-guided processes, microbiological cultures, manual microscopy, manual differentials, etc.).^22^ Tests using research use only (RUO) reagents are included as a subset of in-house developed tests but are not subcategorized as RUOs in our data warehouse, thus precluding separate analysis. EUA assays are included as a subset of FDA tests throughout the manuscript, as they were subject to review and authorization by the FDA under Section 564 of the Federal Food, Drug, and Cosmetic Act.^23^ For clarity, individual assay names were shortened throughout the manuscript to reflect the analyte (and specimen type, as applicable).

Under a separate IRB exempt protocol (University of Utah, #00161484), the number of scanned external test reports categorized as ‘genetic test results’ in our electronic health system (Epic; Verona, WI) in 2021was also retrieved. These results represent assays requested by providers directly to third-party laboratories.

### Design

The total and proportional number of ordered tests and unique assay types were tabulated according to compliance category. These values were then analyzed by patient admission status, ordering location, and ordering department’s medical specialty. The frequency distributions of the most commonly ordered LDTs were then analyzed overall and for each specialty. Data analysis was conducted using Stata 17 (Stata Corp, College Station, TX), Excel 365 (Microsoft: Redmond, CA), and SigmaPlot 14 (Systat: San Jose, CA).

## RESULTS

Providers within our health system ordered 3,020,260 tests through our laboratories in 2021. Of these, 3,332 (0.1%) were for send out tests performed by outside laboratories. As these did not have compliance categories available for review, they were excluded from further analysis. An additional 5,572 scanned test reports categorized as ‘genetic test results’ were identified in our hospital electronic health record system in 2021, representing tests ordered directly by providers to third-party laboratories. Among these, 1,223 had corresponding referral documentation in our laboratory information system that enabled categorization by compliance category. These are analyzed separately, however, as they are not included in our original dataset of test orders.

Of the remaining 3,016,928 tests, 2,831,489 (93.9%) were for FDA assays, 116,583 (3.9%) were for LDTs, and 68,856 (2.3%) were for standard methods (**Table 1**). These test orders were performed using a total of 1,954 distinct assays. Of these, 983 (50.3%) were FDA assays, 880 (45.0%) were LDTs, and 91 (4.7%) were standard methods. Among the FDA tests ordered, 24,385 (0.8% of total orders) were for EUA assays, with more than 99.9% of these orders for SARS-CoV-2 testing. Among the LDT orders, 6,301 were for modified FDA assays (0.2% of total orders), which included 49 unique assays. A change in the specimen type, preservative, or collection device was the reason for the modification of the majority of the assays (40 of 49 assays representing 5,068 of the 6,301 modified FDA tests ordered).

**Table 1:** Categorization of Tests by Regulatory Status.

|  | **Volume of Tests Ordered** | **Distinct Assays** |
| --- | --- | --- |
| **FDA Assays** | **2,831,489 (93.9%)** | **983 (50.3%)** |
| FDA | 2,807,104 (93.0%) | 977 (50.0%) |
| EUA | 24,385 (0.8%) | 4 (0.2%) |
| **LDT Assays** | **116,583 (3.9%)** | **880 (45.0%)** |
| LDT | 110,282 (3.7%) | 831 (42.5%) |
| Modified FDA | 6,301 (0.2%) | 49 (2.5%) |
| **Standard Methods** | **68,856 (2.3%)** | **91 (4.7%)** |
| **Total** | **3,016,928** | **1,954** |

Abbreviations: EUA, emergency use authorization; FDA, Food and Drug Administration; LDT, laboratory-developed test

Test volume and regulatory status were then evaluated by the ordering location and setting (**Table 2**). Overall, total test order volumes were higher at UH compared to the cancer center (2,559,594 and 457,334, respectively). Across these two settings, more tests were ordered in the outpatient (2,207,630, 73.2%; combined) than in the inpatient (809,298, 26.8%; combined) locations. The odd ratio of an exposure to either an LDT or standard test order versus an FDA test order (cancer institute versus UH) was 2.07 (95% CI: 2.01-2.09; P=0.001).

**Table 2.** Test Volumes by Location, Setting, and Regulatory Status.

|  | **FDA** | **LDT** | **Standard** | **Total** |
| --- | --- | --- | --- | --- |
| **University of Utah Health** | **2,422,142 (94.6%)** | **91,133 (3.6%)** | **46,319 (1.8%)** | **2,559,594** |
| Inpatient | 611,060 (95.9%) | 13,255 (2.1%) | 12,911 (2.0%) | 637,226 |
| Outpatient | 1,811,082 (94.2%) | 77,878 (4.1%) | 33,408 (1.7%) | 1,922,368 |
| **Cancer Center** | **409,347 (89.5%)** | **25,450 (5.6%)** | **22,537 (4.9%)** | **457,334** |
| Inpatient | 154,602 (89.8%) | 6,698 (3.9%) | 10,772 (6.3%) | 172,072 |
| Outpatient | 254,745 (89.3%) | 18,752 (6.6%) | 11,765 (4.1%) | 285,262 |
| **Total** | **2,831,489 (93.9%)** | **116,583 (3.9%)** | **68,856 (2.3%)** | **3,016,928** |

Abbreviations: FDA, Food and Drug Administration; LDT, laboratory-developed test

The most commonly ordered LDTs and standard assays across the health system were then evaluated. Ninety percent of the LDT order volume was represented by 167 assays (19.0% of the total number of LDT assays). These assays used predominately four methodologies, including mass spectrometry (60 assays), various immunoassay techniques (33 assays), several nucleic acid amplification techniques (27 assays), and flow cytometry (11 assays) (see **eFigure 1** in the Supplement). The remaining 36 assays used 11 different methodologies. The 20 most frequently ordered LDTs accounted for 53.3% of the total LTD volume (62,095 orders) (**Table 3**). Among the twenty most frequently ordered LDT assays, 12 utilized mass spectrometry, including assays that measure drugs/therapeutics, hormones, vitamins, and trace elements. Seven of the most frequently ordered LDT assays are used in the clinical care of immunocompromised individuals, or in the setting of transplantation, for the detection of tacrolimus, cytomegalovirus (CMV) viral load, CD4 lymphocyte subset, Epstein–Barr virus (EBV) viral load, BK virus viral load, cyclosporin A, and everolimus. Two of the most frequently ordered LDT assays are used for the diagnosis and monitoring of hematopoietic neoplasms (leukemia/lymphoma phenotyping and chromosome analysis). Eight standard assays accounted for 95% of the total standard test volume (**Table 4**).

**Table 3.** Most Frequently Ordered Laboratory-Developed Tests.

| **Test Name** | **Specimen** | **Method** | **Test Volume** | **% LDT Volume** |
| --- | --- | --- | --- | --- |
| Tacrolimus | WB | MS | 12,662 | 10.9% |
| Cytomegalovirus, viral load | P | RT-PCR | 5,226 | 4.5% |
| Estradiol | S, P | MS | 4,527 | 3.9% |
| Leukemia/lymphoma phenotyping | WB | FC | 4,410 | 3.8% |
| Targeted drug profile | U | EIA, MS | 3,394 | 2.9% |
| CD4 lymphocyte subset | WB | FC | 3,373 | 2.9% |
| Vitamin B1 | WB | MS | 3,222 | 2.8% |
| Zinc | S, P | MS | 2,858 | 2.5% |
| Copper | S, P | MS | 2,635 | 2.3% |
| Epstein-Barr virus, viral load | S, P | RT-PCR | 2,584 | 2.2% |
| Progesterone | S, P | MS | 2,279 | 2.0% |
| Vitamin A | S, P | HPLC / UV | 2,144 | 1.8% |
| Chromosome analysis | BM | GB | 1,954 | 1.7% |
| Testosterone, total | S, P | MS | 1,822 | 1.6% |
| Vitamin B6 | S, P | MS | 1,784 | 1.5% |
| Selenium | S, P | MS | 1,618 | 1.4% |
| BK virus, viral load | WB, S, P | RT-PCR | 1,546 | 1.3% |
| Everolimus | WB | MS | 1,445 | 1.2% |
| Albumin, body fluid | BF | SPEC | 1,342 | 1.2% |
| Cyclosporin A | WB | MS | 1,270 | 1.1% |

Abbreviations: LDT, laboratory-developed test; BM, bone marrow; BF, body fluid; FC, flow cytometry; GB, Giemsa Band; HPLC / UV, high-performance liquid chromatography / ultraviolet absorbance; MAA, multi-analyte algorithm; MS, mass spectrometry; IT / Spec, immunoturbidimetry and spectrophotometry; P, plasma; PCR, polymerase chain reaction; RT-PCR, real time polymerase chain reaction; S, serum; U, urine; WB, whole blood

**Table 4.** Most Frequently Ordered Standard Methods.

| **Assay** | **Specimen** | **Method** | **Test Volume** | **% Standard Volume** |
| --- | --- | --- | --- | --- |
| Differential cell count (manual) | WB | M | 25,861 | 37.6% |
| Erythrocyte sedimentation rate | WB | V | 21,209 | 30.8% |
| Urinalysis | U | SPEC | 7,133 | 10.4% |
| Cell count | BF, CSF | M | 5,120 | 7.4% |
| Blood smear with interpretation | WB | M | 2,416 | 3.5% |
| Gram Stain | BF, CSF | M | 1,331 | 1.9% |
| Ova and parasite exam | Stool | M | 1,276 | 1.9% |
| Wet prep, vaginal | G | M | 747 | 1.1% |

Abbreviations: BF, body fluid; CSF, cerebrospinal fluid; G, genital; M, microscopy; SPEC, spectrophotometry; U, urine; V, visual; WB, whole blood

Test volumes according to the ordering location of the medical specialty were then evaluated (**Figure 1**; see **eTable 1** in the Supplement for complete information). Among the orders that could be directly attributed to a specific specialty, internal/family medicine and emergency/intensive care had the highest absolute total numbers of orders (1,044,642 and 456,590, respectively) of which a small percentage were LDT orders (3.2% and 1.4% respectively). LDT orders, as a percentage of total test volumes for the specialty, were the highest in radiology (27.0%), infectious disease (10.4%), and neurology (8.9%). **eTable 2** in the Supplement shows the most common LDTs by medical specialty. Each specialty (except dermatology) included at least one of the top 20 most common LDTs.

**Figure 1.**
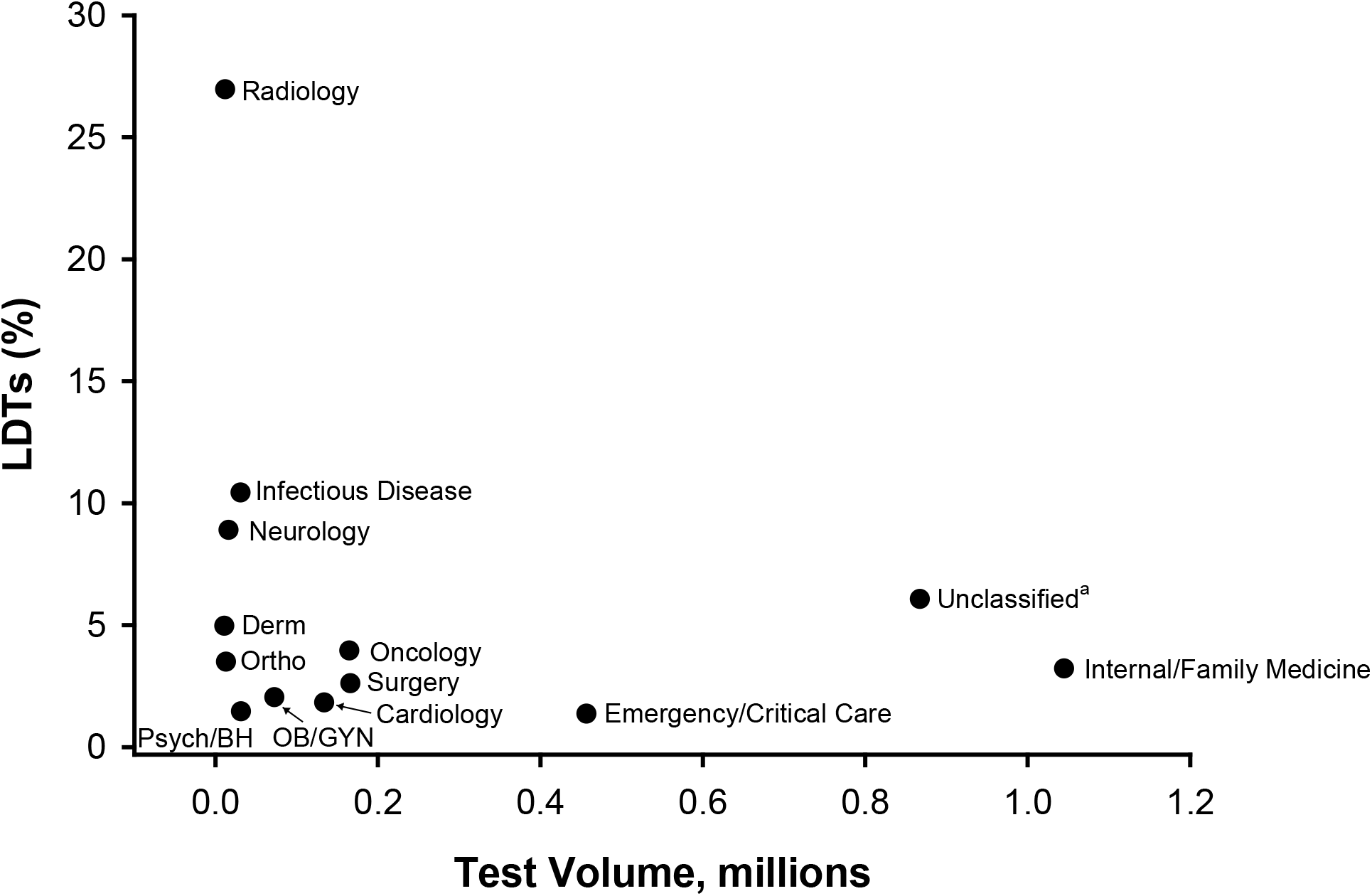
Test Volume Versus Percent Laboratory-Developed Tests (LDTs) by Medical Specialty. Total volume includes FDA assays, LDTs, and standard tests. See Table S1, Supplementary Appendix for complete information Abbreviations: Derm, dermatology; LDT, laboratory-developed test; Psych/BH, psychiatry/behavioral health; OB/GYN, obstetrics and gynecology; Ortho, orthopedics ^a^Unclassified includes orders collected at walk-in outpatient phlebotomy locations

Orthopedics, radiology, and oncology ordered proportionally more standard methods than other specialties (14%, 12.8%, and 6.3%, respectively; **eTable 2** in the Supplement). In orthopedics, the standard methods ordered were primarily erythrocyte sedimentation rates (80.2%) and body fluid cell counts (19.0%); in radiology, the most frequently ordered standard methods were body fluid and CSF cell counts (89.4%); in oncology, the most frequently ordered standard methods were differential cell counts (62.2%) and urine pH (23.2%) (data not shown).

The frequency of orders for all non-infectious disease molecular tests was then evaluated. In our dataset of test orders, a total of 143 different assays were identified for this category, six of which are FDA cleared/approved and included only 62 orders. The remaining 137 were LDT assays accounting for 10,233 orders, representing 8.8% of the total LDT volume and 0.3% of the total order volume (data not shown). Among the most frequently ordered tests in this subclassification were chromosome analysis (1,954 orders), myeloid malignancy panel by NGS (1,138 orders), and quantitative assay for BCR-ABL1 major (p210) fusion forms (839 orders), all of which are used for the diagnosis, prognostic determination, and monitoring of hematopoietic neoplasms. Analysis of the additional 1,223 genetic test reports for assays ordered by providers directly to outside laboratories 9 (for which we had sufficient information to assess compliance category), 137 (11.2%) were for FDA assays and 1,086 (88.8%) were for LDTs.

## DISCUSSION

The present study demonstrated that LDTs account for a relatively low percentage of all diagnostic tests ordered in an academic hospital and outpatient system (3.9% of tests ordered). Furthermore, a relatively small number of LDT assays accounted for the majority of the volume of LDTs ordered (167 assays, 90% of LDT order volume). If standard tests were classified as LDTs, then the combined overall percentage of non-FDA cleared/approved/authorized tests would be 6.1% of all diagnostic tests ordered.

Analysis of our data identified some common themes. First, assays that utilize mass spectrometry are the most common and most frequently ordered LDTs. While clinical mass spectrometers are classified by the FDA as class I (low risk) and exempt from pre-market review,^24^ many of these analytes are currently classified as class II (moderate risk) and would likely require a 510K submission for an assay kit that was commercially distributed by a manufacturer.^25^ While a mass spectrometry assay for the quantitative determination of 25-OH-vitamin D was cleared by the FDA through the De Novo pathway in 2017 and classified as class II (exempt)^26^, for some analytes, including vitamin B_1_, vitamin B_6_, zinc, and copper, there are currently no regulations specifying their risk classification.

The second theme identified was that several of the most frequently ordered LDTs were assays used in the clinical management of immunosuppression and transplantation. These assays are routinely used in evidence-based practice guidelines in patient populations.^27^ While there are FDA-cleared immunoassays for tacrolimus and everolimus and viral load assays for CMV, EBV and BK virus, there appears to be unmet clinical needs that are not addressed by these assays, including non-approved specimen types and workflow modifications.

The third theme we identified in our data analysis was the need for modification of an existing assay for testing alternative specimen types. For example, commonly ordered LDTs in dermatology included PCR testing for herpes simplex virus and Varicella–Zoster virus, for which there are no FDA assays approved for testing specimens other than urogenital or anogenital skin lesions. Because the assays use an alternative specimen type (i.e., a swab from a non-urogenital or non-anogenital lesion), this makes the assay an LDT under current regulatory structures. The same is true for some of the commonly ordered LDTs in emergency medicine (e.g., body fluid today protein, glucose, and lactate dehydrogenase). VALID Act submission requirements related to test modifications due to source and/or specimen type could burden clinical laboratories with significant additional regulatory requirements and costs.^28^ More broadly, given the common themes we identified, it is essential that any impact on overall healthcare costs are considered in the context of regulatory reform efforts.^29^

The present study showed that across specialties, NGS and other non-infectious disease molecular testing are currently not currently among the most commonly ordered methodologies in our health system. This testing made up less than 0.3% of the total testing volume and only 8.8% of the LDT test volume. The regulation of molecular diagnostics has been a particular focus of reform efforts in both the U.S. and the European Union.^30^ NGS and genomic testing, particularly for use in oncology and prenatal screening, are often referenced as justifications in support of diagnostic regulatory reform efforts.^31^ Ultimately, the potential burden and cost of regulatory reform efforts may have a disproportionate impact on the ability to develop and sustain assays that are not frequently ordered.

The differentiation between LDTs and standard tests has not received much attention in diagnostic reform discussions. For example, in the current draft of the VALID Act, “manual tests” are exempt from certain regulatory requirements, but only if they are not considered high-risk and if no component or reagent of such tests is introduced into interstate commerce.^32^ While pathologist interpretations of immunohistochemistry or in situ hybridization may qualify as manual testing, manufacturers of reagents used for manual tests may be subject to new regulatory oversight requirements under the VALID Act.

Limitations of the present study include the fact that data from only one health network were available for the analysis of LDT volumes. Additionally, the presence of a national reference laboratory as part of the university health system may also have contributed to more LDTs being available for ordering than at other institutions. If this were the case, the present study’s findings may over-represent LDT orders versus those placed at other institutions. Alternatively, the present results may represent the combined proportion of LDTs that a large health system would either perform in house or send to referral laboratories. Finally, the present report cannot exclude the possibility of other types of uncategorized scanned test reports in the electronic health record from third-party laboratories.

The observation that only a small percentage of total ordered tests were LDTs and that a relatively small proportion of LDT assays made up the vast majority of the LDTs ordered has practical implications for the potential impact of diagnostic reform efforts on clinical laboratories. For example, an increase in regulatory costs associated with low-volume, low-margin tests could make ongoing clinical offerings unsustainable in certain settings. Regulatory reform efforts should consider all approaches to ensuring the most appropriate and cost-effective patient care is available.

## Data Availability

Research data are not shared.

## ACKNOWLEDGEMENTS

Authors Jenna Rychert, PhD, Robert L. Schmidt, MD, PhD, MBA, and Jonathan R. Genzen, MD, PhD all contributed to the design and analysis of the study and the drafting of the manuscript. Authors Jonathan Genzen and Jenna Rychert had full access to all the data in the study and take responsibility for the integrity of the data and the accuracy of the data analysis. Jonathan Genzen disclosed role as chief medical officer at ARUP Laboratories, and role as principal investigator for contract research to ARUP Laboratories from Fujirebio Diagnostics. Jenna Rychert and Robert Schmidt had no potential conflicts of interest to disclose. The authors acknowledge the assistance of Jason Goodfellow, BS and Michael J. Thompson at ARUP Laboratories for assistance in data retrieval. The authors have no sources of research funding or support to disclose.

