## Supplement for "Laboratory-Developed Test Orders in an Academic Health System"

### Table of Contents

- **eFigure 1**. Volume of LDT Orders by Assay Method Category
- **eTable 1**. Test Volumes and Regulatory Status by Medical Specialty
- **eTable 2**. Common Laboratory-Developed Test Orders by Medical Specialty

**eFigure 1. Volume of LDT Orders by Assay Method Category**

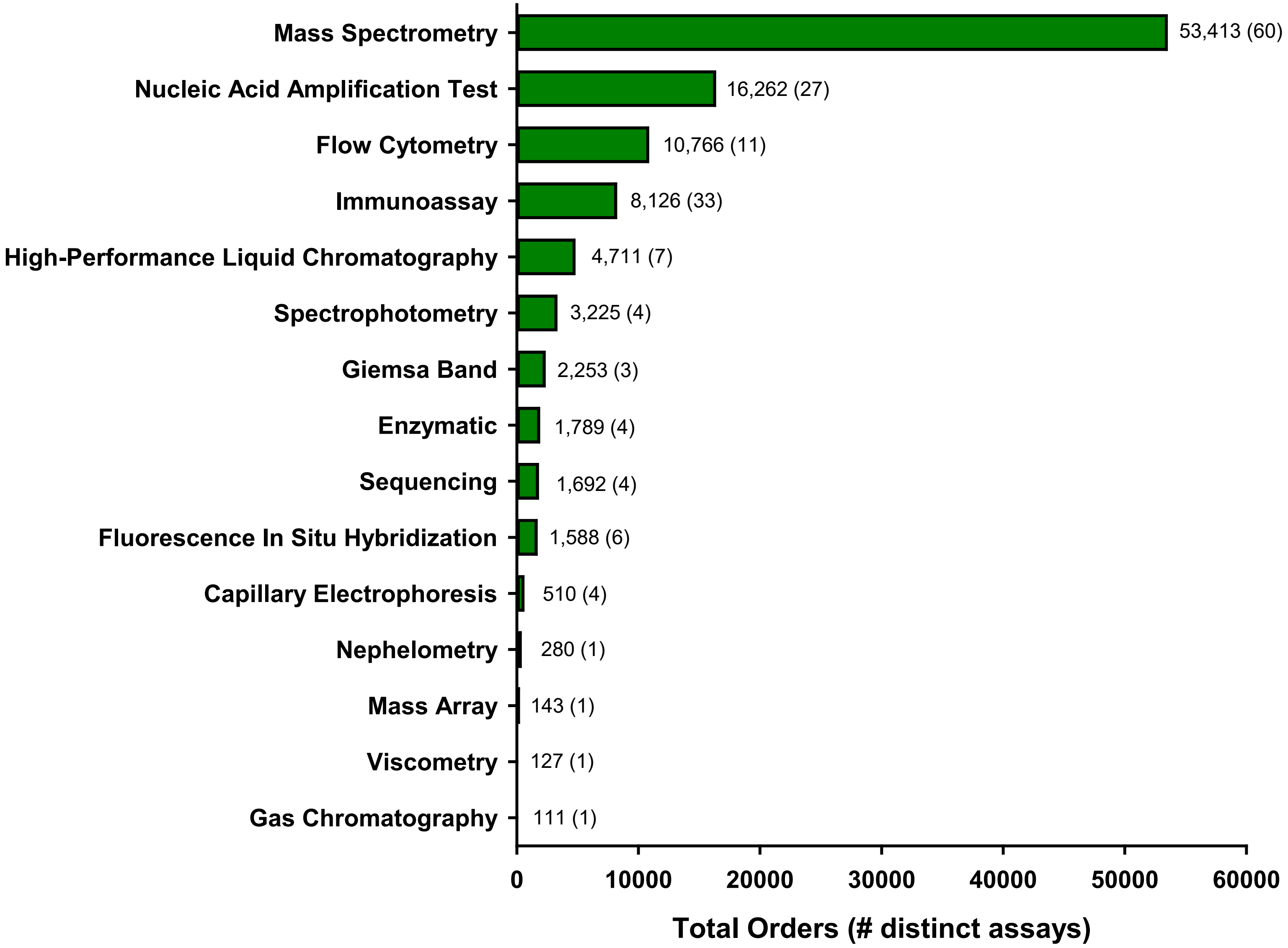

### eTable 1. Test Volumes and Regulatory Status by Medical Specialty

| | FDA | LDT | Standard | Total Volume |
|---|---|---|---|---|
| **Cardiology** | 129,995 (97.4%) | 2,444 (1.8%) | 1,089 (0.8%) | 133,528 |
| **Dermatology** | 9,891 (93.5%) | 526 (5.0%) | 157 (1.5%) | 10,574 |
| **Emergency / intensive care** | 440,148 (96.4%) | 6,280 (1.4%) | 10,162 (2.2%) | 456,590 |
| **Infectious disease** | 27,166 (88.6%) | 3,201 (10.4%) | 309 (1.0%) | 30,676 |
| **Internal / family medicine** | 986,813 (94.5%) | 33,683 (3.2%) | 24,146 (2.3%) | 1,044,642 |
| **Neurology** | 14,103 (89.0%) | 1,411 (8.9%) | 335 (2.1%) | 15,849 |
| **OB/GYN** | 70,140 (97.1%) | 1,478 (2.0%) | 594 (0.8%) | 72,212 |
| **Oncology** | 147,628 (89.8%) | 6,518 (4.0%) | 10,332 (6.3%) | 164,478 |
| **Orthopedics** | 10,439 (82.5%) | 443 (3.5%) | 1,766 (14.0%) | 12,648 |
| **Psychiatry / mental health** | 30,688 (98.2%) | 459 (1.5%) | 93 (0.3%) | 31,240 |
| **Radiology** | 6,951 (60.2%) | 3,112 (27.0%) | 1,478 (12.8%) | 11,541 |
| **Surgery** | 157,320 (94.8%) | 4,355 (2.6%) | 4,241 (2.6%) | 165,916 |
| **Unclassified[a]** | 800,207 (92.3%) | 52,673 (6.1%) | 14,154 (1.6%) | 867,034 |
| **Total** | **2,831,489 (93.9%)** | **116,583 (3.9%)** | **68,856 (2.3%)** | **3,016,928** |

Abbreviations: FDA, Food and Drug Administration; LDT, laboratory-developed test
[a]Includes orders collected at walk-in outpatient phlebotomy locations

**eTable 2. Common Laboratory-Developed Test Orders by Medical Specialty**

| LDT | Specimen | Method | % |
|---|---|---|---|
| **Infectious disease** | | | |
| CD4 lymphocyte subset | WB | FC[1] | 85.4 |
| HLA-B5701 | WB | PCR | 5.6 |
| HIV-1 drug resistance | P | SEQ | 1.9 |
| Histoplasma antigen | S | EIA[1] | 0.7 |
| Vitamin $B_6$ | S, P | MS | 0.6 |
| Histoplasma galactomannan antigen | U | EIA[1] | 0.6 |
| Ureaplasma/Mycoplasma | G | PCR | 0.3 |
| Liver fibrosis, viral | WB+S+P | MAA | 0.3 |
| Hepatitis C genotype | S, P | SEQ | 0.3 |
| Cytomegalovirus viral load | P | PCR | 0.2 |
| **Neurology** | | | |
| Vitamin $B_6$ | S, P | MS | 20.4 |
| Vitamin $B_1$ | WB | MS | 12.3 |
| Copper | S, P | MS | 7.8 |
| Vitamin E | S, P | HPLC | 6.3 |
| Lacosamide | S, P | MS | 4.7 |
| Methylmalonic acid | S, P | MS | 4.6 |
| Oxcarbazepine or Metabolite (MHD) | S, P | MS | 4.4 |
| Clobazam | S, P | MS | 3.1 |
| B-Cell CD20 | S, P | FC | 2.9 |
| Muscle-specific kinase antibody | S | RIA | 2.3 |
| **Internal / family medicine** | | | |
| Targeted drug profile | U | EIA, MS | 7.0 |
| Leukemia/lymphoma phenotyping | WB | FC | 6.0 |
| Cytomegalovirus viral load | P | PCR | 5.1 |
| Tacrolimus | WB | MS | 4.4 |
| Chromosome analysis | BM | GB | 4.3 |
| Epstein-Barr virus, viral load | S, P | PCR | 4.3 |
| Myeloid mutation panel | WB, BM, T | SEQ | 2.5 |
| Estradiol | S, P | MS | 2.5 |
| Lead, capillary | WB | MS | 2.5 |
| Testosterone, total | S, P | MS | 1.7 |

| LDT | Specimen | Method | % |
|---|---|---|---|
| **Dermatology** | | | |
| Varicella-zoster virus | S, P, BF, T, SW | PCR | 18.3 |
| HSV 1/2 subtyping | S, P, BF, T, SW | PCR | 16.2 |
| Thiopurine methyltransferase | RBC | MS | 4.9 |
| Kappa/lambda | T | IHC | 3.4 |
| SOX-10 | T | IHC | 3.0 |
| CD163 | T | IHC | 2.5 |
| Epstein-Barr virus | T | ISH | 2.1 |
| Diphtheria & Tetanus Antibodies, IgG | S | EIA | 2.1 |
| Streptococcus pneumoniae IgG | S | MBA | 2.1 |
| Herpes Simplex Virus (HSV) Types I/II | T | IHC | 2.1 |
| **Orthopedics** | | | |
| Chromium | S | MS | 23.9 |
| Cobalt | S | MS | 23.3 |
| Nicotine and metabolites | S, P | MS | 18.5 |
| Targeted drug profile | U | EIA, MS | 5.0 |
| Targeted drug screen | S, P | EIA | 4.5 |
| HLA-B27 | WB | FC | 4.3 |
| Testosterone, bioavailable & total | S, P | MS | 2.5 |
| Tetrahydrocannabinol | S, P | MS | 1.8 |
| Amphetamines | S, P | MS | 1.6 |
| Estradiol | S, P | MS | 1.6 |
| **Oncology** | | | |
| Tacrolimus | WB | MS | 12.0 |
| Leukemia/lymphoma phenotyping | WB | FC | 10.7 |
| Cytomegalovirus viral load | P | PCR | 8.0 |
| Epstein-Barr virus, viral load | S, P | PCR | 5.8 |
| Chromosome analysis | BM | GB | 3.0 |
| Myeloid mutation panel | WB, BM, T | SEQ | 2.3 |
| Creatinine | BF[1] | SPEC | 2.3 |
| Total protein | BF[1] | SPEC | 2.0 |
| Lactate dehydrogenase | BF[1] | ENZ | 2.0 |
| Amylase | BF[1] | ENZ | 1.7 |

*Continued on next page*

| LDT | Specimen | Method | % |
|---|---|---|---|
| **Obstetrics / gynecology** | | | |
| Progesterone | S, P | MS | 25.6 |
| Amphetamines | U | MS | 11.4 |
| Fentanyl | U | MS | 10.8 |
| Opiates | U | MS | 8.9 |
| Ureaplasma/Mycoplasma | G | PCR | 6.1 |
| Maternal quadruple screen | S | CIA[1] | 5.6 |
| Testosterone, total | S, P | MS | 2.1 |
| Testosterone, free | S, P | MS | 2.1 |
| Fetal Hemoglobin | WB | FC | 2.1 |
| Anti-mullerian Hormone | S | EIA | 2.0 |
| **Psychiatry / mental health** | | | |
| Serotonin | WB | HPLC | 14.8 |
| Clozapine | S, P | MS | 13.9 |
| Tetrahydrocannabinol metabolite | U | MS | 8.7 |
| Targeted drug profile | U | EIA, MS | 4.8 |
| Amphetamines | U | MS | 4.6 |
| Vitamin B1 | WB | MS | 2.8 |
| Targeted drug screen | S, P | EIA | 2.6 |
| Fragile X (FMR1) | WB | PCR, CE | 2.2 |
| Arsenic, Blood | WB | MS | 2.2 |
| Oxcarbazepine or Metabolite (MHD) | S, P | MS | 2.2 |
| **Surgery** | | | |
| Tacrolimus | WB | MS | 26.8 |
| Cytomegalovirus viral load | P | PCR | 10.9 |
| Epstein-Barr virus, viral load | S, P | PCR | 8.7 |
| Leukemia/lymphoma phenotyping | WB | FC | 6.7 |
| Targeted drug screen | S, P | EIA | 2.5 |
| Nicotine and metabolites | S, P | MS | 2.4 |
| Phosphatidylethanol | WB | MS | 2.2 |
| Cyclosporin A | WB | MS | 1.9 |
| Nicotine | U | MS | 1.9 |
| Everolimus | WB | MS | 1.8 |

| LDT | Specimen | Method | % |
|---|---|---|---|
| **Cardiology** | | | |
| Tacrolimus | WB | MS | 38.3 |
| CD3 lymphocyte subset | WB | FC | 6.2 |
| Cytomegalovirus viral load | P | PCR | 4.1 |
| Cyclosporin A | WB | MS | 2.6 |
| Everolimus | WB | MS | 2.2 |
| Heparin-induced thrombocytopenia antibody | S | EIA | 1.8 |
| Sirolimus | WB | MS | 1.7 |
| Pneumocystis jirovecii | R | PCR | 1.6 |
| Targeted drug screen | S, P | EIA | 1.5 |
| Chlamydia pneumoniae | R | PCR | 1.4 |
| **Emergency / intensive care** | | | |
| Targeted drug screen | S, P | EIA | 5.9 |
| Leukemia/lymphoma phenotyping | WB | FC | 4.6 |
| Vitamin $B_1$ | WB | MS | 3.7 |
| Total protein | BF[1] | SPEC | 3.7 |
| HSV 1/2 subtyping | S, P, BF, T, SW | PCR | 3.5 |
| Glucose | BF[1] | ENZ | 3.3 |
| Lactate dehydrogenase | BF[1] | ENZ | 2.5 |
| Albumin | BF[1] | SPEC | 2.5 |
| Cytomegalovirus viral load | P | PCR | 2.3 |
| Varicella-zoster virus | S, P, BF, T, SW | PCR | 2.2 |
| **Radiology** | | | |
| Albumin | BF[1] | SPEC | 26.4 |
| Leukemia/lymphoma phenotyping | WB | FC | 22.6 |
| Chromosome analysis | BM | GB | 8.9 |
| Total protein | BF[1] | SPEC | 4.9 |
| Multiple myeloma MRD | BM | FC | 4.5 |
| Multiple myeloma | BM, WB | FISH | 4.4 |
| MGMT promoter methylation | T | PCR, MA, MS | 2.9 |
| Myeloid mutation panel | WB, BM, T | SEQ | 2.6 |
| Lactate dehydrogenase | BF[1] | ENZ | 2.1 |
| Glucose | BF[1] | ENZ | 1.9 |

Abbreviations: Ab, antibody; Ag, antigen; BF, body fluid; CC, cell culture; CIA, chemiluminescent immunoassay; ENZ, enzymatic; FC, flow cytometry; GB, Giemsa Band; IHC, immunohistochemistry; ISH, in situ hybridization; MA, mass array; MAA, multi-analyte algorithm; MGMT, methylguanine-DNA methyltransferase; MHD, monohydroxy carbamazepine; MRD, minimal residual disease; MS, mass spectrometry; PCR, polymerase chain reaction; P, plasma; S, serum; SEQ, sequencing; SPEC, spectrophotometry; T, tissue; U, urine; UMB, umbilical; WB, whole blood; G, genital; RBC, red blood cells; RIA, radioimmunoassay; SW, swab. [1] Modified FDA
